# Taste loss as a distinct symptom of COVID-19: A systematic review and meta-analysis

**DOI:** 10.1101/2021.10.09.21264771

**Authors:** Mackenzie E. Hannum, Riley J. Koch, Vicente A. Ramirez, Sarah S. Marks, Aurora K. Toskala, Riley D. Herriman, Cailu Lin, Paule V. Joseph, Danielle R. Reed

## Abstract

Chemosensory scientists have been skeptical that reports of COVID-19 taste loss are genuine, in part because before COVID-19, taste loss was rare and often confused with smell loss. Therefore, to establish the predicted prevalence rate of taste loss in COVID-19 patients, we conducted a systematic review and meta-analysis of 376 papers published in 2020–2021, with 241 meeting all inclusion criteria. Additionally, we explored how methodological differences (direct vs. self-report measures) may affect these estimates. We hypothesized that direct prevalence measures of taste loss would be the most valid because they avoid the taste/smell confusion of self-report. The meta-analysis showed that, among 138,897 COVID-19-positive patients, 39.2% reported taste dysfunction (95% CI: 35.34–43.12%), and the prevalence estimates were slightly but not significantly higher from studies using direct (n = 18) versus self-report (n = 223) methodologies (Q = 0.57, df = 1, p = 0.45). Generally, males reported lower rates of taste loss than did females and taste loss was highest in middle-aged groups. Thus, taste loss is a bona fide symptom COVID-19, meriting further research into the most appropriate direct methods to measure it and its underlying mechanisms.

## Introduction

The novel coronavirus (COVID-19), a respiratory infection caused by severe acute respiratory syndrome coronavirus-2 (SARS-CoV-2), was first identified in Wuhan, China, and has since spread throughout the world. When the World Health Organization first declared this a pandemic in March 2020, researchers and clinicians were not yet aware that the virus affected individuals’ senses of smell and taste, but these symptoms soon became apparent via patient reports. As a result of COVID-19, affected people can experience chemosensory dysfunction in a variety of ways, including complete loss of smell or taste (anosmia or ageusia, respectively), partial loss of smell or taste (hyposmia or hypogeusia), and/or a distorted sense of smell or taste (e.g., parosmia, dysgeusia). These chemosensory dysfunctions can be distressing to the affected individuals and can last for extended times, with some patients experiencing resolution within a few weeks to a month (Lee et al., 2020b; Gerkin et al., 2021) and others with symptoms for 6 months or longer (Blomberg et al., 2021).

Previous meta-analyses examined smell and taste loss in COVID-19 patients, but often with a focus on onset and duration (Santos et al., 2021) or recovery (Boscutti et al., 2021) of chemosensory symptoms. Many focused only on smell loss (Hannum et al., 2020; Pang et al., 2020; Rocke et al., 2020) or general neurological symptoms (Abdullahi et al., 2020; Favas et al., 2020; Mair et al., 2021; Yassin et al., 2021). Very few continued to evaluate articles published in 2021 and often capped reviewing articles 6-10 months after March 2020, when the pandemic was declared, limiting the number of articles included (ranging from 5 to 59 articles total). Therefore, we decided to conduct a more comprehensive analysis, spanning a year and a half, to ensure a fuller coverage of the available research.

Additionally, taste loss is often neglected in research compared to smell loss, as there is a common notion that taste loss is not as “real” as smell loss. Some claim taste loss is indistinguishable from smell loss (Le Bon et al., 2021) or is confused with smell loss (Deems et al., 1991), specifically with retronasal smell perception (Hintschich et al., 2020a). For the general population, loss of taste can be difficult to distinguish from smell loss. Therefore, it may be difficult to know, based on self-report measures alone, whether or not a participant truly lost their sense of taste.

Thus, many chemosensory researchers may attribute the taste loss phenomena seen in the current reports of COVID-19-positive patients to deficiencies of self-report or subjective measures of taste loss. Therefore, we conducted a systematic review and meta-analysis to estimate the true prevalence of taste loss in COVID-19 patients across a wide sample of studies (n=241) and to evaluate effects of major methodological differences in data collection. In particular, we compared overall findings on taste loss as determined by individual taste tests (herein referred to as direct tests) with self-reports without direct sensory testing. We hypothesized that direct methodologies would support the presence of taste loss as a distinct symptom, and direct measures might even be higher than self-report despite the possible inflation of self-reported taste loss which is exacerbated by smell loss.

Currently scientists are using both direct and self-report measures to examine chemosensory dysfunction, with self-report far more common due to the pandemic restrictions, e.g., sensory laboratories where direct testing is often conducted are closed. For taste, direct tests include standardized and non-standardized tests that contain various sweet, salty, and sometimes bitter and sour stimuli given to participants via solutions, drops, strips, or sprays (Cao et al., 2021; Singer-Cornelius et al., 2021). Non-standardized direct taste measures created to study COVID-19-related taste dysfunction include solution-based tests, often prepared at home by participants (Vaira et al., 2020f). Self-report measures include interviews with researchers and clinicians, electronic health records as well as surveys administered over the phone, online, or in person.

To understand taste loss as a symptom of COVID-19, we conducted a large systematic review and meta-analysis, examining how it has been measured (direct vs. self-report) and how the measurement type can affect prevalence rates. We tested the hypothesis that direct measures are at least as sensitive as self-report measures and would confirm taste loss as a distinct symptom and not merely misattributed smell loss.

## Methods

### Article Selection

This systematic review and meta-analysis followed the Preferred Reporting Items for Systematic Reviews and Meta-Analyses guidelines (Moher et al., 2009). Articles were selected via searches on Pubmed/Medline and Google Scholar, using the keyword “COVID-19” with “taste”, “smell”, and/or “olfaction”, as well as “gustatory”. Literature retrieval began on May 15, 2020, and concluded on June 1, 2021, resulting in 377 articles in total.

Initial screening of the articles included reading the titles and abstracts to assess their relevance. Articles with an abstract that reported chemosensory dysfunction in COVID-19-infected individuals were included in the systematic review (*n* = 376). Next, at least two authors read the articles initially deemed relevant, to evaluate whether they fit the inclusion criteria: reporting positive COVID-19 tests, written in the English language, and lack of population bias. COVID-19 must have been confirmed via nasopharyngeal swab, reverse transcription polymerase chain reaction (RT-PCR), or assessment by physician or other medical personnel. The articles were then evaluated on whether they reported taste loss data specifically. In total, 135 articles were excluded based on such criteria as not evaluating taste loss, recruiting participants with chemosensory dysfunction, not testing for COVID-19, and presenting overlapping data (see Figure 1). In total, 241 articles were included in the final meta-analysis (corresponding citations described in “Included Articles” section at the end of the paper).

**Figure 1.**
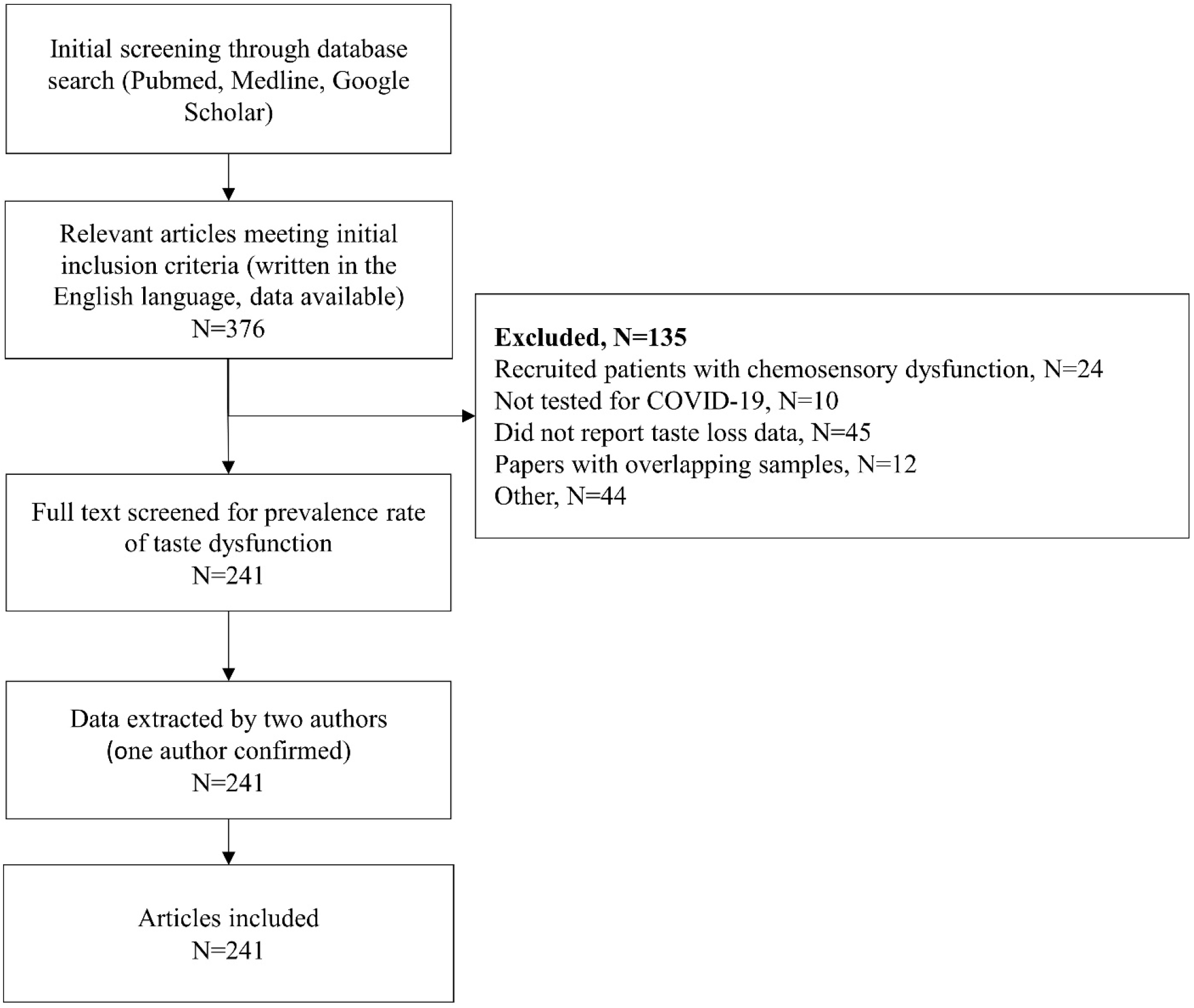
CONSORT flow diagram demonstrating the article selection process for this systematic review and meta-analysis.

### Data Extraction

We extracted from each article either the number or percentage of patients with taste dysfunction due to a SARS-CoV-2 infection. The prevalence of taste loss reported in each article was calculated by dividing the reported number of participants with taste loss as a symptom by the total number of COVID-19-positive participants. Additionally, measures of taste loss were labeled as “self-report” or “direct” to identify the method used to evaluate participants. Self-report measures included reported loss of taste via surveys, interviews, and electronic medical health records. Most articles (n = 223) used self-report methods. Table 1 summarizes the studies that used direct measures (n = 18), comprising actual taste tests administered either at home (Adamczyk et al., 2020; Hintschich et al., 2020a; Petrocelli et al., 2020b; Cao et al., 2021; Singer-Cornelius et al., 2021), at a testing facility (Altin et al., 2020; Bidkar et al., 2020a; Mazzatenta et al., 2020; Ramteke et al., 2020; Vaira et al., 2020a; Le Bon et al., 2021; Niklassen et al., 2021a; Salcan et al., 2021) or both in home and in a hospital environment (Vaira et al., 2020b, 2020c, 2020d, 2020f). Vaira et al. [2020e] had an unknown testing location. Many of the measures consisted of solution-based tests measuring four basic taste sensations: sweet, sour, salty, and bitter.

**Table 1.** Overview of the direct approaches to assess taste loss in participants.

| *Test* | *Article* | *Test Method* | *Test Objective* | *Taste Quality Measured* |  |  |  |  |
| --- | --- | --- | --- | --- | --- | --- | --- | --- |
|  |  |  |  | *Salty* | *Sweet* | *Sour* | *Bitter* | *Umami* |
| Four-solution test | Vaira et al., 2020a–2020f<br>Petrocelli et al., 2020<sup>A</sup> | 1 mL of each solution, plus deionized water as control, placed on the participant's tongue via cotton swab. Quarantined patients prepared their own solutions. | Identification | ☒ | ☒ | ☒ | ☒ | ☐ |
| Four-solution test & Taste Strips | Altin et al., 2020<br>Salcan et al., 2021 | Participants swallowed and identified the solution. Next, paper strips dipped into each solution and placed on the participant's tongue. | Identification<br>Duration | ☒ | ☒ | ☒ | ☒ | ☒ |
| Two-solution test (1) | Bidkar et al., 2020 | Two drops (2 mL) of each solution placed on the participant's tongue via pipette. | Identification | ☒ | ☒ | ☐ | ☐ | ☐ |
| Two-solution test (2) | Mazzatenta et al., 2020 | 200 µl of each solution dropped onto participant's tongue. | Detection | ☒ | ☒ | ☐ | ☐ | ☒ |
| Taste Strips by BMG<sup>B</sup> | Hintschich et al., 2020<br>Singer-Cornelius et al., 2021<br>Le Bon et al., 2021<br>Niklassen et al., 2021<sup>C</sup> | Taste strips (provided by Burghart Messtechnik GmbH) placed on the participant's tongue. | Identification and threshold concentration | ☒ | ☒ | ☒ | ☒ | ☐ |
| Taste sprays | Niklassen et al., 2021<sup>C</sup> | Each solution sprayed onto participant's tongue. | Identification | ☒ | ☒ | ☒ | ☒ | ☐ |
| Tastant capsules | Adamczyk et al., 2020 | Each of ten 0.33 mL gelatin capsules (one tasteless and nine with tastant) dissolved on participant's tongue. | Description | ☒ | ☒ | ☒ | ☒ | ☐ |
| Brief Self-Administered Waterless Empirical Taste Test (SA-WETT)<sup>B</sup> | Cao et al., 2021 | 27 disposable plastic strips that contain dried solutions of each taste (and some tasteless strips) self-placed on participant's tongue. | Identification | ☒ | ☒ | ☒ | ☒ | ☒ |
| Chemosensory Test by India Protocol | Ramteke et al., 2020 | Authors used coconut oil, chocolates, and flavored milk to test smell and taste function. No further details are provided. | N/A | N/A |  |  |  |  |

<sup>A</sup> These studies used a sweet solution concentration that is double what was used in the other four-solution tests.
<sup>B</sup> Validated test.
<sup>C</sup> Niklassen et al. (2021) used both Taste Strips and taste sprays with participants.

Although some authors reported exclusively smell loss or taste loss, many reported on both senses. Thus, it was necessary to include additional coding options for when authors reported the symptoms in tandem (e.g., “loss of smell or taste”). Therefore, articles were labeled as “taste only”, “smell and/or taste”, “smell and taste”, “smell or taste”, and “smell and/or taste; taste only” (in the event both values were reported, the numbers were summed), depending on how these symptoms were phrased in the article.

We also extracted demographic characteristics of each study, including the population mean and/or median age, sex (expressed as percentage of males in the population), and country of origin (for the geographic distribution of the study populations, see Supplementary Figure 1 (S1)).

Four authors performed the initial reading of full texts and the data extraction from the studies (R.D.H., A.K.T., S.S.M., R.J.K.). Two authors confirmed this information and resolved any inconsistencies (M.E.H., D.R.R.). Differences were resolved through a discussion and renewed consensus on the proposed solution from all authors who read that specific article.

### Risk-of-Bias Assessment

We used a risk-of-bias assessment from Hoy et al. (2012) to examine the articles selected for the meta-analysis. The assessment contained nine questions, outlined in Supplementary Materials (see Supplementary Table (S2)). Responses were scored as 1 (no) or 0 (yes), with summary scores of low (0-3), moderate (4-6), and high (7-9). Two authors completed the risk-of-bias assessment of each article using the checklist developed by Hoy et al. (2012), as described and adapted by Tong et al. (2020) (S.S.M. and A.K.T.). One author resolved any discrepancies (M.E.H.).

### Statistical Analysis

The meta-analysis was conducted using the meta package in R (Schwarzer et al., 2019). Generalized linear mixed models were used for the meta-analysis, as recommended for the analysis of binary outcomes and proportions (Bakbergenuly and Kulinskaya, 2018; Schwarzer et al., 2019). Heterogeneity (e.g., between study variance) was assessed using Cochran’s Q, I^2^, and tau squared (τ^2^). We concluded there was evidence for heterogeneity when the p-value for Cochran’s Q was less than 0.05 and if the I^2^ was greater than 50% (Higgins and Thompson, 2002). Tau squared (τ^2^), a measure of between study variance, was estimated using the maximum likelihood approach and has no associated test-statistic.

An overall pooled prevalence estimates were computed and reported for a random-effects model with the parameters described for all 241 studies. While both fixed and random-effect models were computed, excess the heterogeneity among our studies (see the Results section) suggested that unmeasured effects contribute to variance in our data more than would be expected from sampling error. The extreme diversity of taste loss as reported in individual studies (0 to 93.4%) suggested that the random-effects model was more appropriate.

Subgroup analysis was performed for studied employing direct methods (n = 18) and self-report methods (n = 223) to assess taste function in COVID-19–positive individuals. Additionally, the average age of participants and their sex (percentage of male subjects in each study) were included as covariates in univariate mixed-regression models. Subgroup analysis for age was arbitrarily categorized into five groupings: adolescents (0-18 years old), young adults (19-35 years old), middle-aged adults (36-50 years old), older adults (51-65 years old), and elderly adults (65+ years old). Finally, studies that used direct tests were separated, and subgroup analysis was performed for each type of collection methodology: solution based (N=12), strip based (N=5), and other (N=1). For continuous variables (e.g., age), the transformed beta coefficients are reported. The transformation for the generalized linear mixed models uses a logit transformation for each proportion, so the models are interpreted as the log(odds) or log(p/1 – p), where p is equal to the prevalence for each study. Back-transformation into the prevalence estimates were done for subgroup analysis (e.g., age).

All statistical analyses were performed using R 4.0.5 (R Core, 2021) and RStudio 1.4.1106 (RStudio Team, 2020). Visualization of the meta-analysis is displayed as an orchard plot adapted from Nakagawa et al. (2020). The R scripts and compiled data used for this analysis are available without restriction at GitHub (https://github.com/vramirez4/COVID19-TasteLoss).

## Results

### Risk-of-Bias Assessment

Each of the 241 articles included in this meta-analysis were reviewed for risk of bias. Among these articles, none had a high risk of bias, 142 studies had a moderate risk, and 99 studies had a low risk (see Supplementary Table S2 for the full assessment).

### Prevalence of taste loss in COVID-19-positive patients

Among the 241 studies, the sample sizes ranged from 11 to over 40,000 patients with COVID-19. The number of cases of taste loss per study ranged from 0 to 4,668, with raw prevalence estimates ranging from 0% to 93.4%. Collectively, the meta-analysis included 138,785 patients who tested positive for COVID-19. Of these, 32,918 patients had some form of taste loss after infection with SARS-CoV-2. Heterogeneity among the prevalence estimates across all studies (n = 241) yielded a significant Cochran’s Q (Q = 29388.75, degrees of freedom [df] = 240, P < 0.001), and an I^2^ estimate of 99.2%, and τ^2^ estimate of 1.5. The pooled estimate for taste loss prevalence in COVID-19-positive patients following meta-analysis for the overall cohort was 36.90% (95% CI: 33.27–40.69%).

### Effect of methodology (direct vs. self-report) on prevalence estimate

We employed a subgroup analysis to determine the effect of direct versus self-report approaches on taste loss prevalence (see Figure 2). Eighteen studies used direct methods to assess taste loss, comprising 2,240 COVID-19 patients, with 1,092 reported cases of taste loss. Per study, the prevalence of taste loss ranged from 0% to 84% among COVID-19-positive patients. For studies using direct approaches, the pooled estimate of the prevalence for the random-effect model was 42.0% (95% CI: 30.0-55.0%). Cochran’s Q was significant (Q = 187.93, df = 17, P < 0.001), and an I^2^ of 91.0% was obtained, confirming heterogeneity of the data collected via direct measures. The τ^2^ for the direct methodologies was 1.2.

**Figure 2.**
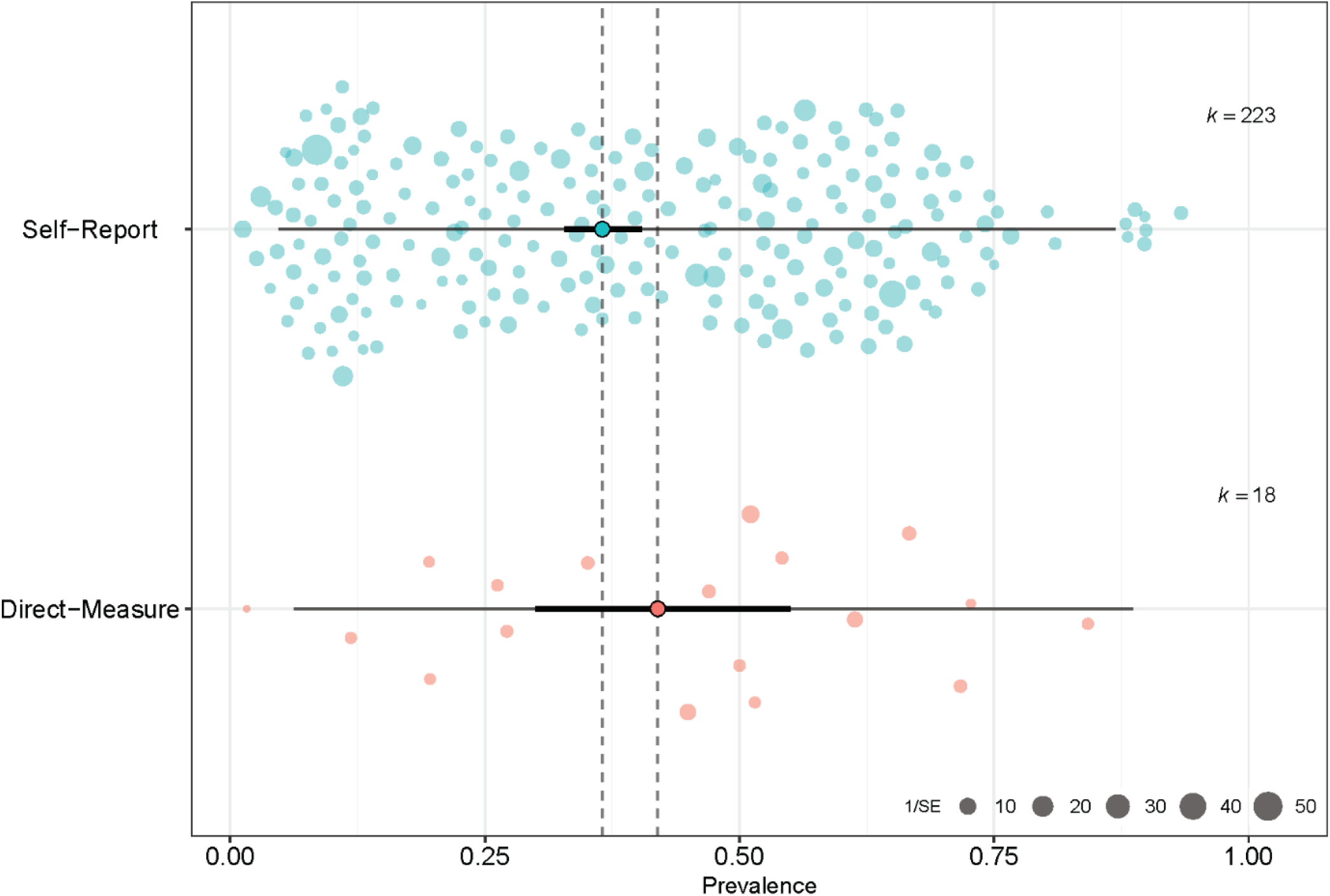
Orchard plot of taste loss and COVID-19. The point estimate of the pooled prevalence (trunk) is represented by the bold turquoise or pink dot. The confidence interval of the pooled prevalence estimate (branch) is represented by the bold black line, and the prediction interval (twig) is represented by the thin black line. Individual prevalence estimates from each study are represented by the scattered colored points (slightly transparent circles). Each point is scaled by the precision of the point estimate of prevalence for each study, i.e., inverse of the standard error.

A total of 223 studies used self-report methods (e.g., questionnaire, interview), comprising 136,545 subjects, with 31,826 cases of taste loss. The reported prevalence of taste loss ranged from 1% to 93% per study. The pooled estimate of the prevalence under the random-effect models was 36.53% (95% CI: 32.8%-40.5%). Similar to the direct subgroup, Cochran’s Q was significant (Q = 29188.14, df = 222, P < 0.001), and the I^2^ value was 99.2%, confirming that the studies differed in prevalence across the self-report studies. The τ^2^ for self-reported studies was 1.5.

A test of heterogeneity between groups was employed using Cochran’s Q, which revealed that the differences between methodologies were not statistically significant (Q = 0.66, df = 1, p = 0.4157) under the random-effects model. While our analysis showed that the prevalence of taste loss was higher when measured directly than by self-report, there was no significant effect of measurement method on the prevalence estimates of taste loss.

### Effect of age and sex on taste loss prevalence

Additional analyses were undertaken to assess the effect of age and sex. Univariate mixed models for each covariate revealed that both age and sex had significant effects on the prevalence of taste loss. After categorizing each study by mean age group, we conducted the meta-analysis for those 210 studies that reported the ages of the participants (see Table 2). Among this subset of studied the pooled prevalence was 37.16% and demonstrated significant heterogeneity with (Q=33943.46, df = 209, p<0.001), I^2^ = 98.9, and τ^2^ =1.5. The prevalence estimates per age category ranged from 11% in studies with average ages younger than 18 years to 44% in studies with average ages between 36 and 50 years. Heterogeneity in these studies was high (I^2^ = 96.4-98.9%) both within groups (Q = 14106.77, df = 205, p<0.0001) and between groups (Q = 32.19, df = 4, p<0.0001). The results demonstrated that both the youngest and oldest age groups report the lowest prevalence of taste loss, while age groups between 18 and 65 years had pooled estimates ranging from 32% to 44%, with the highest in the middle-age (36-50 years) group. Similarly, it appeared that age accounted for some of the overall heterogeneity among some groups as measured by reductions in τ^2^ among studies who contained mostly adolescent and elderly individuals.

**Table 2.**
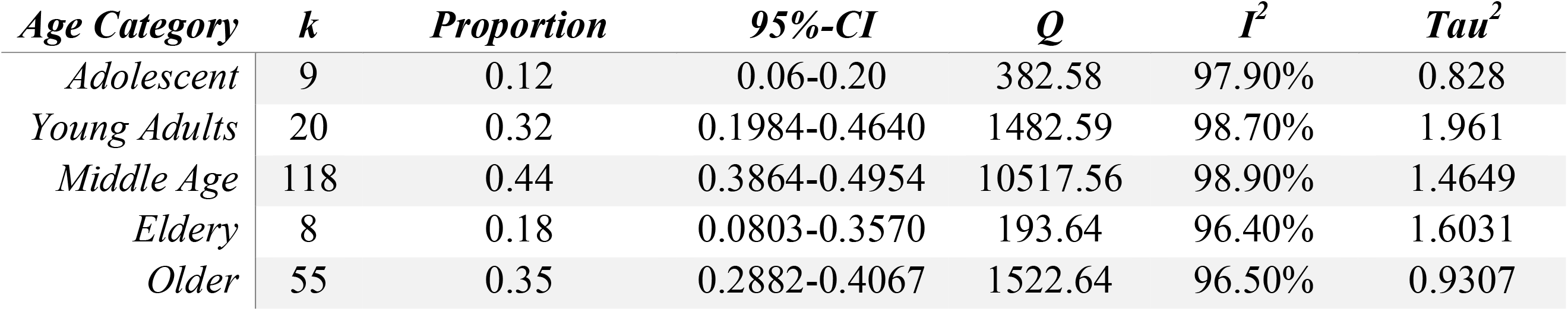
Random-effect estimate of age group on COVID-19 taste loss prevalence using generalized linear mixed models.

| *Age Category* | *k* | *Proportion* | *95%-CI* | *Q* | *I<sup>2</sup>* | *Tau<sup>2</sup>* |
| --- | --- | --- | --- | --- | --- | --- |
| *Adolescent* | 9 | 0.12 | 0.06-0.20 | 382.58 | 97.90% | 0.828 |
| *Young Adults* | 20 | 0.32 | 0.1984-0.4640 | 1482.59 | 98.70% | 1.961 |
| *Middle Age* | 118 | 0.44 | 0.3864-0.4954 | 10517.56 | 98.90% | 1.4649 |
| *Eldery* | 8 | 0.18 | 0.0803-0.3570 | 193.64 | 96.40% | 1.6031 |
| *Older* | 55 | 0.35 | 0.2882-0.4067 | 1522.64 | 96.50% | 0.9307 |

The effects of sex were similarly examined. A univariate mixed model using percentage of males in each study as a covariate found an effect of β = −0.0296 (p < 0.001) for each percent increase. Overall, the higher the percentage of males in a study, the lower the prevalence of taste loss.

### Effect of type of direct approach on taste loss prevalence

When we compared the types of direct report test (e.g., solution-based test, taste strip-based test), we found a significant difference in prevalence rates (see Table 3). We classified studies into three categories: taste strip testing (n=5), taste solution testing (n = 12), and “other” (n = 1) for methods that do not employ either solution or strips. Pooled prevalence for solution-based tests was 54.25% (95% CI: 45.01-63.19%) and for taste strips was 24.63% (95% CI: 13.10-41.46%). There was reduced heterogeneity in the subgroups compared to the overall meta-analysis: solution-based testing, I^2^ = 88.8%; and strip-based testing, I^2^ = 76.8%. Measurements of τ^2^, were 0.3807 and 0.5991 for solution-based testing and strip-based testing respectively. There was significant heterogeneity between our pooled estimates (Q = 8.68, df = 2, p = 0.01). Together, our results demonstrate that studies using solution-based taste tests, on average, result in higher prevalence of taste loss in COVID-19 patients than do studies using strips or other methods.

**Table 3.**
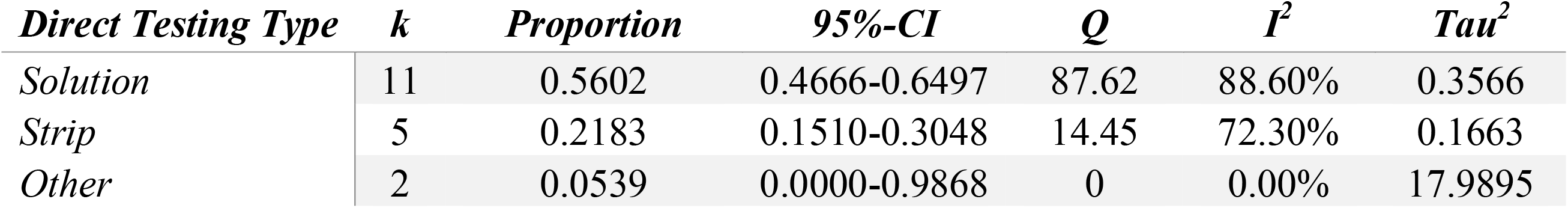
Random-effect estimate of direct testing type on COVID-19 taste loss prevalence using generalized linear mixed models.

## Discussion

Despite the occurrence of true taste loss in a variety of diseases such as cancer (Nolden et al., 2019), as well as in the general population (Rawal et al., 2016), taste loss has often been confused with smell loss (Le Bon et al., 2021). However, the current coronavirus pandemic suggests that in reality, taste loss is its own unique feature of the illness. The present meta-analysis found an overall taste loss prevalence of 37 % among 138,897 COVID-19-positive participants, which aligns with other meta-analyses of taste loss prevalence, ranging from 38% (Agyeman et al., 2020) to 49% (Hajikhani et al., 2020). This high prevalence is not due to confusion with smell loss because direct taste measures yield similar (or even slightly higher) prevalence than self-report. Therefore self-reports of taste loss appear to be valid among people with COVID-19 as they are among other groups (Jang et al., 2021).

The COVID-19 pandemic created an urgent need for direct taste measures suitable for the pandemic research environment, e.g., home testing, and researchers were innovative, but each developed their own method which makes it difficult to compare results (see Table 1). However, despite the differences in methods, we can draw one general conclusion, which is that the form of the tastants matters (taste solutions are better than taste strips) but this general conclusion must be tentative given that the forms of delivery were not compared directly using the same approach, e.g., thresholds or identification.

The present meta-analysis found that around 4 in every 10 COVID-19 patients experience taste loss. We also found age and sex effects: females experienced higher rates of taste loss than males, aligning with other meta-analyses reporting a similar effect (von Bartheld et al., 2020; Amorim Dos Santos et al., 2021; Saniasiaya et al., 2021). Females may be more susceptible to taste loss because they are in general are more sensitive than males and have more sensory capacity to lose. Additionally, we found that COVID-19 associated taste loss peaks in middle age aligning with the general consensus across other COVID-19 meta-analyses (Agyeman et al., 2020; von Bartheld et al., 2020). Why the youngest and oldest groups report less taste loss than do middle-age adults is not currently known.

Although COVID-19 has intensified awareness of taste loss and furthered chemosensory research, scientists are still unsure of the biological mechanisms behind this symptom. The amount of SARS-CoV-2 virus in saliva is positively related to loss of taste: the more virus, the more taste loss (Huang et al., 2021; Taziki Balajelini et al., 2021), although this observation is controversial (Jain et al., 2020). Taste cells may be attacked directly by the virus because studies of expression patterns for ACE2, the receptor protein known to transport the SARS-CoV-2 virus into cells, and for TMPRSS2, the protein essential for processing the SARS-CoV-2 spike protein, showed both are expressed in the supporting cells of taste bud (Sakaguchi et al., 2020; Huang et al., 2021), including taste receptor cells themselves, at least in one patient (Doyle et al., 2021). There may also be direct effects on the brain that contribute to taste loss (Douaud et al., 2021).

### Limitations and Future Research

In many of the included articles, clinicians and researchers collected self-reported taste loss information in tandem with smell loss (e.g., participants responding yes to “Loss of taste and smell” on a symptom screener), which can confound the results. Therefore, we explored any differences in how taste loss was collected, reflecting how it was reported in the articles (e.g., “smell and taste” vs. “taste only”), and found no significant impact on the prevalence rate (see Supplementary Materials S3). This result indicates that taste loss is a common and pervasive symptom of COVID-19. Additionally, far more articles in this meta-analysis used self-report tests (n = 223) than direct tests (n = 18). This disparity may have prevented us from capturing significant differences between the two methods.

Nearly all of the articles included in this meta-analysis were nonspecific to different tastes, instead summarizing scores across multiple stimuli (e.g., sweet and salty) and reporting taste loss as a whole (though in one study participants self-reported taste-specific dysfunction: salt taste loss, 29.3%; sweet taste, 25.9%; general taste, 34.5% [El Kady et al., 2021]). However, specific taste sensitivities can be difficult to assess via self-report. Members of the general population, untrained in chemosensory science, may have difficulties identifying whether or not they truly lost a specific taste. Therefore, it is important to use direct measures to distinguish dysfunctions of specific tastes.

There are clear opportunities for advancements of standardized direct taste tests to measure taste loss. Of the 18 studies that used direct tests in general, only five used standardized tests, representing just 2.06% of the 242 studies examined in this systematic review: Taste Strips by Burghart Messtechnik (Hintschich et al., 2020a; Le Bon et al., 2021; Niklassen et al., 2021a; Singer-Cornelius et al., 2021) and the Brief Self-Administered Waterless Empirical Taste Test (SA-WETT) by Sensonics International (Cao et al., 2021). Three other studies using direct tests were examined during the systematic review but were excluded for not meeting the inclusion criteria: reporting on a case study (Lee and Lee, 2020), recruiting patients with chemosensory dysfunction (Le Bon et al., 2020), and not reporting the required taste loss prevalence data (Huart et al., 2020). Among these three studies, Lee and Lee (2020) used a non-standardized direct test, and Huart et al. (2020) and Le Bon et al. (2020) used the Taste Strips – although this represents a missed opportunity to analyze more studies that used standardized tests, including these two articles would have increased the rate of standardized test use among all 241 studies to just 2.89% (N=7).

As of September 2021, 220 million individuals have been infected with SARS-CoV-2 virus, with large number of recovered individuals with persistent symptoms included taste loss. It is well documented that disease-related or age-related or chemosensory loss have profound effects on an individual’s quality of life. Unlike other disorders such as vision and hearing for which preventative and screening guidelines exist (United States Preventive Services Taskforce), they are not available for taste and smell disorders. The COVID-19 pandemic further highlighted this existing gap, lack of standardized measures and clinical guidelines for screening, assessing, and monitoring the taste system making it difficult for clinicians to track progression of disease. Assessment of taste function in patients with and without confirmed COVID-19 needs to become standard of practice for clinicians. This is particularly important for at least two reasons: 1) having baseline measures help clinicians assess trends over time, and 2) given the interrelatedness between the sense of smell and taste, objective measures of taste collected during clinical assessments may help dissociate whether it is a smell or taste problem. For patients who report changes in taste function during screening questionnaires, full testing with standardized objective chemosensory tools should be performed. It is critical that clinicians are aware that most patients with chemosensory dysfunction complain of taste alterations, therefore, a closer inquiry of patient’s reports regarding the specific taste quality (i.e., sweet, bitter, sour, and salt, fat) affected is important to further distinguish between taste and perception of flavor.

## Conclusion

The COVID-19 pandemic demanded an urgent response from scientists and clinicians, who have been working to understand the novel virus and the symptoms it inflicts. Of the many unique features of this virus, smell and taste dysfunctions are among the most prominent. Yet as taste loss joined smell loss as a more prolific topic in scientific literature, many initially speculated that taste loss rates were overestimated due to confusion between taste and smell in self-reports. However, our meta-analysis found a prevalence rate for taste loss of 36.9% among 138,897 COVID-19-positive individuals (95% CI: 33.27–40.69%), supported by direct methods, reflecting the validity of this distinct symptom. Dysfunction in the sense of taste was, and still is, a difficult reality for millions of people affected by the virus and merits further research to fully understand the mechanisms behind this phenomenon and how to properly assess and address it. Among the population of 138,897 COVID-19-positive individuals included in this meta-analysis, only 257 of them, across five separate studies, were assessed using standardized taste tests. Future research should include the development of fast and accurate taste tests, studies that measure taste and smell function separately to dissociate olfacto-gustatory interactions, as well as the employment of standardized taste tests in clinical settings to examine taste dysfunction. In addition, clinical trials are needed to elucidate frequency of screening and age at which to start and stop screening for chemosensory disorders. Finally, more mechanistic studies to understand taste and smell disorders associated with COVID-19 to aid in developing new therapeutic options for patients with long-lasting impairment of their chemical senses.

## Data Availability

All data produced in the present work are contained in the manuscript

## Included Articles

In total, 241 were included in the present systematic review and meta-analysis (Gamper et al., 2012; Liu et al., 2016; Adamczyk et al., 2020; Adedeji et al., 2020; Adorni et al., 2020; Aggarwal et al., 2020; Al-Ani and Acharya, 2020; Altin et al., 2020; Andrews et al., 2020; Anna et al., 2020; Asai et al., 2020; Bastiani et al., 2020; Beltran-Corbellini et al., 2020; Bergquist et al., 2020; Biadsee et al., 2020; Bidkar et al., 2020b; Boscolo-Rizzo et al., 2020a, 2020b, 2020c, 2021; Boudjema et al., 2020; Brandao Neto et al., 2020; Bulgurcu et al., 2020; Calica Utku et al., 2020; Carignan et al., 2020; Chary et al., 2020; Chen et al., 2020; Chiesa-Estomba et al., 2020; Cho et al., 2020; Chung et al., 2020; Cocco et al., 2020; Dawson et al., 2020; Dell’Era et al., 2020; De Maria et al., 2020; Durrani et al., 2020; Elimian et al., 2020; Farah Yusuf Mohamud et al., 2020; Fistera et al., 2020; Fontanet et al., 2020; Freni et al., 2020; Garg et al., 2020; Gelardi et al., 2020; Giacomelli et al., 2020; Gorzkowski et al., 2020; Gozen et al., 2020; GÜner et al., 2020; Gudbjartsson et al., 2020; Guillen Martinez et al., 2020; Haehner et al., 2020; Hintschich et al., 2020b; Horvath et al., 2020; Iversen et al., 2020; Izquierdo-Domínguez et al., 2020; Jain et al., 2020; Jalessi et al., 2020; Kacem et al., 2020; Karadas et al., 2020; Kempker et al., 2020; Kim et al., 2020; Klopfenstein et al., 2020; Konstantinidis et al., 2020; Krishnasamy et al., 2020; Kronborg et al., 2020; Kumar et al., 2020, 2021a, 2021b; Lagi et al., 2020; Lan et al., 2020; Lapostolle et al., 2020; La Torre et al., 2020; Lechien et al., 2020b, 2020c, 2020a, 2021, 2021, 2021; Lechner et al., 2020; Lee et al., 2020a, 2020b, 2021; Levinson et al., 2020; Liang et al., 2020; Liguori et al., 2020; Lindahl et al., 2020; Lombardi et al., 2020, 2021; Luers et al., 2020; Lv et al., 2020; Maechler et al., 2020; Magnavita et al., 2020; Makda et al., 2020; Mao et al., 2020; Martin-Sanz et al., 2020; Mazzatenta et al., 2020; Meini et al., 2020; Menni et al., 2020; Mercante et al., 2020; Merkely et al., 2020; Merza et al., 2020; Moein et al., 2020; Morshed et al., 2020; Murat et al., 2020; Nakagawa et al., 2020, 2020; Nakakubo et al., 2020; Nakanishi et al., 2020; Noh et al., 2020; Otte et al., 2020; Ozcelik Korkmaz et al., 2020; Paderno et al., 2020; Patel et al., 2020; Perlman et al., 2020; Petersen et al., 2020; Petrocelli et al., 2020b, 2020a; Pinna et al., 2020; Printza et al., 2020, 2021; Qiu et al., 2020; Rajkumar et al., 2020; Ramteke et al., 2020, 2020; Rojas-Lechuga et al., 2020; Roland et al., 2020; Romero-Sanchez et al., 2020; Rubel et al., 2020; Sakalli et al., 2020; Salepci et al., 2020; Sayin et al., 2020a, 2020b; Schirinzi et al., 2020; Schmithausen et al., 2020; Seo et al., 2020; Shoer et al., 2020; Sierpinski et al., 2020; Smith et al., 2020; Somekh et al., 2020; Speth et al., 2020; Spinato et al., 2020; Stavem et al., 2020; Sun et al., 2020; Teklu et al., 2020; Tham et al., 2020; Tsivgoulis et al., 2020; Tudrej et al., 2020; Vacchiano et al., 2020; Vaira et al., 2020e, 2020f, 2020b, 2020c, 2020d, 2020a, 2020b; Van Loon et al., 2020; Vena et al., 2020; Villarreal et al., 2020; Wagner et al., 2020; Waterfield et al., 2020; Wee et al., 2020; Weiss et al., 2020; Weng et al., 2020; Yan et al., 2020a, 2020b; Zayet et al., 2020a, 2020b; Zimmerman et al., 2020; Akinbami et al., 2021; Alharbi et al., 2021; AlShakhs et al., 2021; Amano et al., 2021; Arslan et al., 2021; Ashrafi et al., 2021; Bagnasco et al., 2021; Barillari et al., 2021; Besli et al., 2021; Bianco et al., 2021; Breyer et al., 2021; Callejon-Leblic et al., 2021; Cao et al., 2021; Carcamo Garcia et al., 2021; Concheiro-Guisan et al., 2021; Dini et al., 2021; Dixon et al., 2021; El Kady et al., 2021; Fisher et al., 2021; Fleischer et al., 2021; Galluzzi et al., 2021; Gianola et al., 2021; Gibbons et al., 2021; Gonzalez et al., 2021; Gupta et al., 2021a, 2021b; Gurrola et al., 2021; Hijazi et al., 2021; Ismail et al., 2021; Jethani et al., 2021; Kamel et al., 2021; Kandakure et al., 2021; Karaarslan et al., 2021; Karni et al., 2021; Kavaz et al., 2021; Ladoire et al., 2021; Lampl et al., 2021; Makaronidis et al., 2021; Mannan et al., 2021; Moeller et al., 2021; Monti et al., 2021; Morlock et al., 2021; Niklassen et al., 2021b; Ninchritz-Becerra et al., 2021; Nishanth et al., 2021; Noviello et al., 2021; Oda et al., 2021; Omezli and Torul, 2021; O’Keefe et al., 2021; O’Sullivan et al., 2021; Ozcan et al., 2021; Parcha et al., 2021; Perula de Torres et al., 2021; Polat et al., 2021; Rodebaugh et al., 2021; Rousseau et al., 2021; Sahoo et al., 2021; Salcan et al., 2021; Savtale et al., 2021; Sbrana et al., 2021; Schwab et al., 2021; Sehanobish et al., 2021; Singer-Cornelius et al., 2021; Soh et al., 2021; Song et al., 2021a, 2021b; Tarifi et al., 2021; Thakur et al., 2021; Trachootham et al., İşlek and Balcı, 2021; van den Besselaar et al., 2021; Wierdsma et al., 2021; Yadav et al., 2021; Zejda et al., 2021; Zifko et al., 2021; Zou et al., 2021)

## Acknowledgements

We acknowledge Michael G. Tordoff for his assistance with the review of the literature, Sarah Lipson for her initial effort in reading the earlier articles, and Akane Kikuchi for ensuring no article was missed. Additionally, we thank Gary K. Beauchamp for his comments on the manuscript.

