## Supplemental materials for "Taste loss as a distinct symptom of COVID-19: A systematic review and meta-analysis"

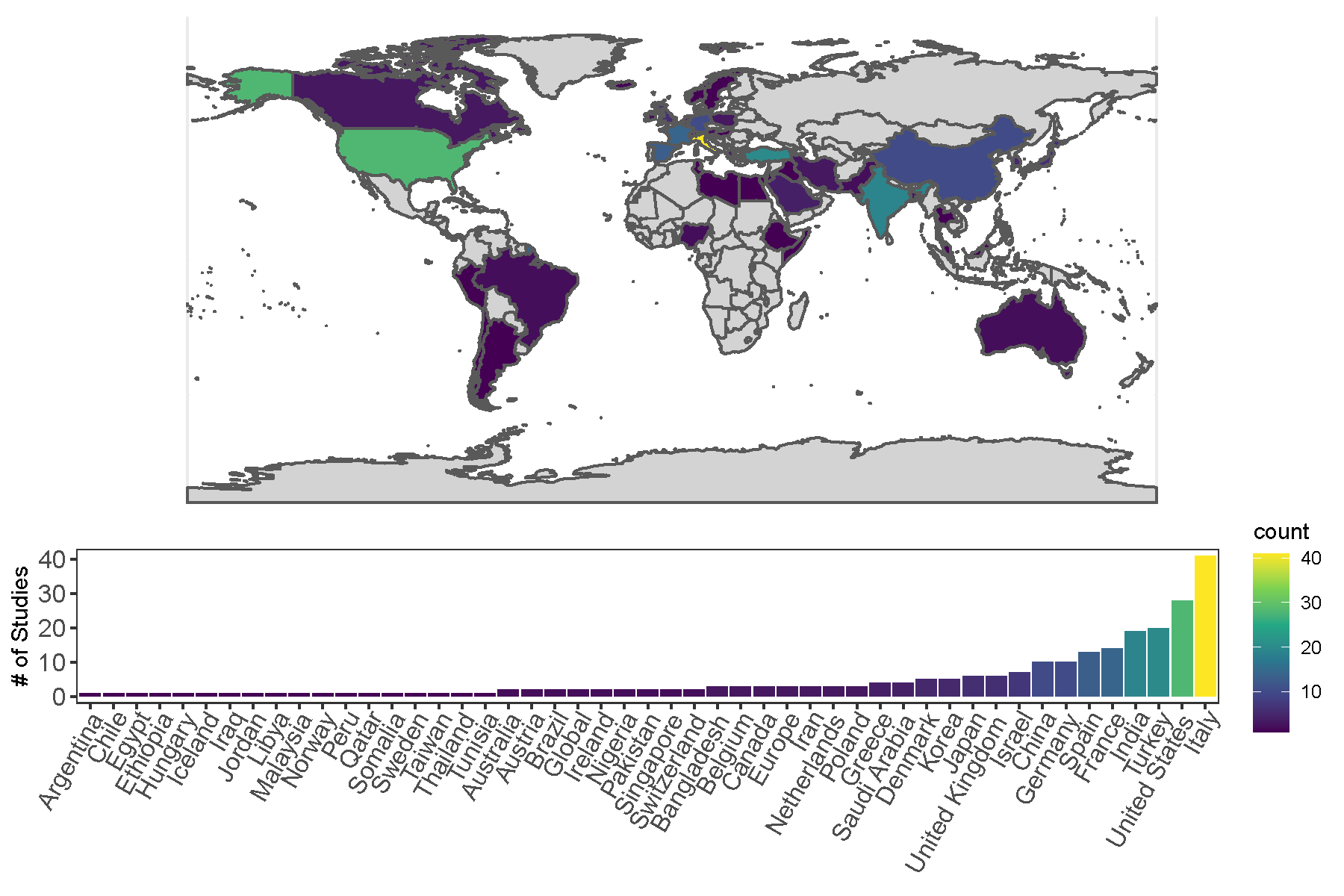

**S1: Supplementary Figure.** Geographic distribution of patient populations included in the present systematic review and meta-analysis. Color gradient corresponds to number of research articles from the country (count legend (far left): yellow – more articles; purple – fewer articles; grey (on map) – no articles).

**S2: Supplementary Table.** Risk-of-bias assessment of selected articles.

| **Article** | **Question^A^** | | | | | | | | | **Overall Risk** | **Risk Category^B^** |
| --- | --- | --- | --- | --- | --- | --- | --- | --- | --- | --- | --- |
|  | **Q1** | **Q2** | **Q3** | **Q4** | **Q5** | **Q6** | **Q7** | **Q8** | **Q9** |  |  |
| **Vaira et al. 1** | 1 | 0 | 1 | 0 | 0 | 0 | 0 | 0 | 0 | 2 | Low |
| **Vaira et al. 2** | 1 | 0 | 1 | 0 | 0 | 0 | 1 | 1 | 0 | 4 | Moderate |
| **Merza et al.** | 1 | 0 | 1 | 0 | 1 | 0 | 1 | 1 | 0 | 5 | Moderate |
| **Haehner et al.** | 0 | 0 | 1 | 0 | 1 | 0 | 1 | 0 | 0 | 3 | Low |
| **Speth et al.** | 0 | 0 | 1 | 0 | 1 | 0 | 1 | 0 | 0 | 3 | Low |
| **De Maria et al.** | 1 | 0 | 1 | 0 | 0 | 0 | 1 | 1 | 0 | 4 | Moderate |
| **Menni et al. 2** | 0 | 0 | 1 | 0 | 0 | 0 | 0 | 0 | 0 | 1 | Low |
| **Yan et al. 1** | 1 | 0 | 1 | 0 | 0 | 0 | 1 | 0 | 0 | 3 | Low |
| **Luers & Rokohl et al.** | 0 | 0 | 1 | 1 | 0 | 0 | 0 | 0 | 0 | 2 | Low |
| **Roland et al.** | 0 | 0 | 1 | 0 | 1 | 0 | 1 | 0 | 0 | 3 | Low |
| **Boscolo‑Rizzo et al.** | 1 | 0 | 1 | 0 | 0 | 0 | 1 | 0 | 0 | 3 | Low |
| **Liu et al.** | 0 | 0 | 1 | 0 | 0 | 0 | 1 | 0 | 0 | 2 | Low |
| **Paderno & Schreiber et al.** | 1 | 0 | 1 | 0 | 1 | 0 | 1 | 0 | 0 | 4 | Moderate |
| **Lee et al.** | 1 | 0 | 1 | 0 | 0 | 0 | 1 | 0 | 0 | 3 | Low |
| **Lechien & Chiesa‐Estomba et al.** | 1 | 0 | 1 | 0 | 0 | 0 | 1 | 0 | 0 | 3 | Low |
| **Gelardi et al.** | 1 | 0 | 1 | 0 | 0 | 0 | 0 | 1 | 0 | 3 | Low |
| **Giacomelli et al.** | 1 | 0 | 1 | 0 | 1 | 0 | 1 | 0 | 0 | 4 | Moderate |
| **Lechien et al. 2** | 1 | 0 | 1 | 1 | 1 | 0 | 1 | 0 | 0 | 5 | Moderate |
| **Shoer & Karady et al.** | 1 | 0 | 1 | 0 | 0 | 0 | 0 | 1 | 0 | 3 | Low |
| **Vaira et al. 3** | 1 | 0 | 1 | 0 | 0 | 0 | 0 | 1 | 0 | 3 | Low |
| **Mao & Wang et al.** | 1 | 0 | 1 | 0 | 0 | 0 | 0 | 1 | 0 | 3 | Low |
| **Spinato et al.** | 1 | 0 | 1 | 1 | 0 | 0 | 1 | 0 | 0 | 4 | Moderate |
| **Beltran-Corbellini et al.** | 1 | 0 | 1 | 1 | 0 | 0 | 0 | 0 | 0 | 3 | Low |
| **Trubiano et al.** | 1 | 0 | 1 | 0 | 0 | 0 | 1 | 0 | 0 | 3 | Low |
| **Moein et al.** | 1 | 0 | 1 | 0 | 1 | 0 | 1 | 1 | 0 | 5 | Moderate |
| **Yan et al. 2** | 1 | 0 | 1 | 0 | 0 | 0 | 0 | 0 | 0 | 2 | Low |
| **Gudbjartsson & Helgason et al.** | 1 | 0 | 1 | 0 | 1 | 0 | 1 | 0 | 0 | 4 | Moderate |
| **Wee et al.** | 0 | 0 | 0 | 1 | 0 | 0 | 1 | 1 | 0 | 3 | Low |
| **Dawson & Rabold et al.** | 1 | 0 | 1 | 0 | 1 | 0 | 1 | 0 | 0 | 4 | Moderate |
| **Noh et al.** | 1 | 0 | 1 | 0 | 1 | 0 | 1 | 0 | 0 | 4 | Moderate |
| **Tudrej et al.** | 1 | 0 | 1 | 0 | 1 | 0 | 1 | 0 | 0 | 4 | Moderate |
| **Sayin et al.** | 1 | 0 | 1 | 0 | 0 | 0 | 1 | 0 | 0 | 3 | Low |
| **Dell'era et al.** | 1 | 0 | 1 | 0 | 0 | 0 | 1 | 1 | 0 | 4 | Moderate |
| **Baidsee & Biadsee et al.** | 1 | 0 | 1 | 0 | 0 | 0 | 1 | 0 | 0 | 3 | Low |
| **Qiu & Cui et al.** | 1 | 0 | 1 | 0 | 0 | 0 | 1 | 0 | 0 | 3 | Low |
| **Izquierdo-Dominguez & Rojas-Lechuga et al.** | 1 | 0 | 1 | 0 | 0 | 0 | 1 | 1 | 0 | 4 | Moderate |
| **Weng et al.** | 1 | 0 | 1 | 0 | 0 | 0 | 1 | 0 | 0 | 3 | Low |
| **Lombardi et al.** | 1 | 0 | 1 | 0 | 1 | 0 | 1 | 0 | 0 | 4 | Moderate |
| **Freni et al.** | 1 | 0 | 1 | 0 | 1 | 0 | 0 | 0 | 0 | 3 | Low |
| **Altin et al.** | 1 | 0 | 1 | 1 | 0 | 0 | 1 | 0 | 0 | 4 | Moderate |
| **Schmithausen & Döhla et al.** | 1 | 0 | 1 | 0 | 0 | 0 | 0 | 0 | 0 | 2 | Low |
| **Durrani et al.** | 0 | 0 | 0 | 0 | 0 | 0 | 1 | 0 | 0 | 1 | Low |
| **Mercante et al.** | 1 | 0 | 1 | 0 | 1 | 0 | 1 | 0 | 0 | 4 | Moderate |
| **Lan et al.** | 1 | 0 | 1 | 0 | 0 | 0 | 1 | 0 | 0 | 3 | Low |
| **Hintschich et al.** | 1 | 0 | 1 | 0 | 1 | 0 | 1 | 0 | 0 | 4 | Moderate |
| **Vacchiano et al.** | 1 | 0 | 1 | 0 | 0 | 0 | 1 | 0 | 0 | 3 | Low |
| **Petrocelli et al.** | 1 | 0 | 1 | 0 | 0 | 0 | 1 | 0 | 0 | 3 | Low |
| **Lechien & Chiesa-Estomba et al. 3** | 1 | 0 | 1 | 0 | 0 | 0 | 0 | 0 | 0 | 2 | Low |
| **Sakalli et al.** | 1 | 0 | 1 | 0 | 0 | 0 | 1 | 1 | 0 | 4 | Moderate |
| **Boscolo‑Rizzo et al. 2** | 0 | 0 | 1 | 0 | 0 | 0 | 1 | 1 | 0 | 3 | Low |
| **Adorni et al.** | 1 | 0 | 1 | 0 | 0 | 0 | 0 | 0 | 0 | 2 | Low |
| **Somekh et al.** | 1 | 0 | 1 | 0 | 1 | 0 | 0 | 0 | 0 | 3 | Low |
| **Chiesa-Estomba & Lecien et al.** | 0 | 0 | 1 | 0 | 0 | 0 | 1 | 0 | 0 | 2 | Low |
| **Güner et al.** | 1 | 0 | 1 | 0 | 0 | 0 | 0 | 0 | 0 | 2 | Low |
| **Martin-Sanz et al.** | 1 | 0 | 1 | 0 | 1 | 0 | 1 | 0 | 0 | 4 | Moderate |
| **Boscolo-Rizzo et al. 2** | 1 | 0 | 1 | 0 | 0 | 0 | 1 | 0 | 0 | 3 | Low |
| **Villarreal et al.** | 1 | 0 | 1 | 0 | 0 | 0 | 1 | 1 | 0 | 4 | Moderate |
| **Nakagawara et al.** | 1 | 0 | 1 | 0 | 0 | 0 | 1 | 0 | 0 | 3 | Low |
| **Wagner & Shweta et al.** | 1 | 0 | 1 | 0 | 1 | 0 | 1 | 0 | 0 | 4 | Moderate |
| **Fontanet et al.** | 1 | 0 | 1 | 0 | 1 | 0 | 1 | 1 | 0 | 5 | Moderate |
| **Kempker et al.** | 1 | 0 | 1 | 1 | 0 | 0 | 1 | 0 | 0 | 4 | Moderate |
| **Chen et al.** | 1 | 0 | 1 | 0 | 0 | 0 | 1 | 0 | 0 | 3 | Low |
| **Gorzkowski et al.** | 1 | 0 | 1 | 0 | 0 | 0 | 1 | 0 | 0 | 3 | Low |
| **Morshed et al.** | 1 | 0 | 1 | 0 | 0 | 0 | 1 | 0 | 0 | 3 | Low |
| **Moein et al. 2** | 1 | 0 | 1 | 0 | 1 | 0 | 1 | 0 | 0 | 4 | Moderate |
| **Cocco et al.** | 1 | 0 | 1 | 0 | 1 | 0 | 0 | 0 | 0 | 3 | Low |
| **Vaira et al. 4** | 1 | 0 | 1 | 0 | 0 | 0 | 1 | 0 | 0 | 3 | Low |
| **Klopfenstein et al. 2** | *1* | 0 | 1 | 0 | 0 | 0 | 1 | 0 | 0 | 3 | Low |
| **Vaira et al. 5** | 1 | 0 | 1 | 0 | 1 | 0 | 1 | 0 | 0 | 4 | Moderate |
| **Cho et al.** | 0 | 0 | 1 | 1 | 1 | 0 | 1 | 0 | 0 | 4 | Moderate |
| **Salepci et al.** | 1 | 0 | 1 | 0 | 0 | 0 | 1 | 1 | 0 | 4 | Moderate |
| **Mohamud et al.** | 1 | 0 | 1 | 0 | 0 | 0 | 1 | 0 | 0 | 3 | Low |
| **Kronborg et al.** | 1 | 0 | 1 | 0 | 1 | 0 | 1 | 0 | 0 | 4 | Moderate |
| **Merkely et al.** | 1 | 0 | 1 | 0 | 1 | 0 | 1 | 0 | 0 | 4 | Moderate |
| **Jalessi et al.** | 1 | 0 | 0 | 1 | 0 | 0 | 1 | 0 | 0 | 3 | Low |
| **Utku et al.** | 1 | 0 | 0 | 0 | 1 | 0 | 1 | 0 | 0 | 3 | Low |
| **Maeshler et al.** | 1 | 0 | 0 | 0 | 0 | 0 | 1 | 0 | 0 | 2 | Low |
| **Chary et al.** | 1 | 0 | 1 | 0 | 1 | 0 | 1 | 0 | 0 | 4 | Moderate |
| **Bergquist et al.** | 1 | 0 | 1 | 0 | 1 | 0 | 1 | 0 | 0 | 4 | Moderate |
| **Al-Ani et al.** | 1 | 0 | 1 | 0 | 1 | 0 | 1 | 0 | 0 | 4 | Moderate |
| **Otte et al.** | 0 | 0 | 1 | 0 | 1 | 0 | 1 | 0 | 0 | 3 | Low |
| **Rojas-Lechuga et al.** | 0 | 0 | 1 | 0 | 0 | 0 | 1 | 0 | 0 | 2 | Low |
| **Lechien et al. 4** | 1 | 0 | 1 | 0 | 1 | 0 | 1 | 1 | 0 | 5 | Moderate |
| **Van Loon et al.** | 1 | 0 | 1 | 0 | 1 | 0 | 1 | 1 | 0 | 5 | Moderate |
| **Neto et al.** | 1 | 0 | 1 | 0 | 0 | 0 | 1 | 0 | 0 | 3 | Low |
| **Vena et al.** | 1 | 0 | 1 | 0 | 0 | 0 | 1 | 0 | 0 | 3 | Low |
| **Weiss et al.** | 1 | 0 | 0 | 0 | 0 | 0 | 1 | 0 | 0 | 2 | Low |
| **Chiesa-Estomba et al.** | 1 | 0 | 1 | 1 | 0 | 0 | 1 | 0 | 0 | 4 | Moderate |
| **Carignan et al.** | 1 | 0 | 1 | 0 | 1 | 0 | 0 | 0 | 0 | 3 | Low |
| **Kim et al.** | 1 | 0 | 1 | 0 | 0 | 0 | 0 | 0 | 0 | 2 | Low |
| **Lee et al. 2** | 1 | 0 | 1 | 0 | 0 | 0 | 1 | 0 | 0 | 3 | Low |
| **Liguori et al.** | 1 | 0 | 1 | 1 | 0 | 0 | 1 | 0 | 0 | 4 | Moderate |
| **Meini et al.** | 1 | 0 | 1 | 0 | 1 | 0 | 1 | 0 | 0 | 4 | Moderate |
| **Sierpiński et al.** | 1 | 0 | 1 | 0 | 0 | 0 | 1 | 1 | 0 | 4 | Moderate |
| **Zayet et al.** | 1 | 0 | 1 | 0 | 0 | 0 | 1 | 0 | 0 | 3 | Low |
| **Aggarwal et al.** | 1 | 0 | 1 | 0 | 1 | 0 | 1 | 0 | 0 | 4 | Moderate |
| **Patel et al.** | 1 | 0 | 1 | 0 | 1 | 0 | 1 | 0 | 0 | 4 | Moderate |
| **Romero-Sanchez et al.** | 1 | 0 | 1 | 1 | 1 | 0 | 1 | 0 | 0 | 5 | Moderate |
| **Karni et al.** | 1 | 0 | 1 | 0 | 1 | 0 | 1 | 0 | 0 | 4 | Moderate |
| **Lindahl et al.** | 1 | 0 | 1 | 0 | 0 | 0 | 0 | 0 | 0 | 2 | Low |
| **Perlman et al.** | 1 | 0 | 1 | 0 | 1 | 0 | 1 | 1 | 0 | 5 | Moderate |
| **Lv et al.** | 1 | 0 | 1 | 0 | 0 | 0 | 1 | 0 | 0 | 3 | Low |
| **Vaira et al. 6** | 1 | 0 | 1 | 0 | 0 | 0 | 1 | 0 | 0 | 3 | Low |
| **Korkmaz et al.** | 1 | 0 | 1 | 0 | 0 | 0 | 0 | 0 | 0 | 2 | Low |
| **Rajkumar et al.** | 1 | 0 | 1 | 0 | 1 | 0 | 1 | 0 | 0 | 4 | Moderate |
| **Ramasamy et al.** | 1 | 0 | 1 | 0 | 0 | 0 | 1 | 0 | 0 | 3 | Low |
| **Nakanishi et al.** | 1 | 0 | 1 | 0 | 0 | 0 | 0 | 0 | 0 | 2 | Low |
| **Konstantinidis et al.** | 1 | 0 | 1 | 0 | 1 | 0 | 1 | 0 | 0 | 4 | Moderate |
| **Makda et al.** | 1 | 0 | 1 | 1 | 1 | 0 | 1 | 0 | 0 | 5 | Moderate |
| **Zimmerman et al.** | 1 | 0 | 1 | 0 | 0 | 0 | 1 | 0 | 0 | 3 | Low |
| **Bidkar et al.** | 1 | 0 | 1 | 0 | 1 | 0 | 1 | 0 | 0 | 4 | Moderate |
| **Seo et al.** | 1 | 0 | 1 | 0 | 0 | 0 | 1 | 1 | 0 | 4 | Moderate |
| **Alshami et al.** | 1 | 0 | 1 | 0 | 0 | 0 | 0 | 0 | 0 | 2 | Low |
| **Smith et al.** | 0 | 0 | 1 | 0 | 1 | 0 | 1 | 0 | 0 | 3 | Low |
| **Schirinzi et al.** | 1 | 0 | 1 | 0 | 1 | 0 | 1 | 0 | 0 | 4 | Moderate |
| **Tham et al.** | 1 | 0 | 1 | 1 | 0 | 0 | 1 | 0 | 0 | 4 | Moderate |
| **Waterfield et al.** | 1 | 0 | 1 | 0 | 1 | 0 | 1 | 0 | 0 | 4 | Moderate |
| **Murat et al.** | 1 | 0 | 1 | 0 | 0 | 0 | 1 | 0 | 0 | 3 | Low |
| **Adedeji et al.** | 1 | 0 | 1 | 0 | 1 | 0 | 1 | 0 | 0 | 4 | Moderate |
| **Jain et al.** | 1 | 0 | 1 | 1 | 1 | 0 | 1 | 0 | 0 | 5 | Moderate |
| **Kacem et al.** | 1 | 0 | 1 | 0 | 0 | 0 | 1 | 1 | 0 | 4 | Moderate |
| **Krishnasamy et al.** | 0 | 0 | 1 | 0 | 0 | 0 | 0 | 0 | 0 | 1 | Low |
| **Nakakubo et al.** | 1 | 0 | 1 | 0 | 1 | 0 | 1 | 0 | 0 | 4 | Moderate |
| **Rubel et al.** | 1 | 0 | 1 | 0 | 1 | 0 | 1 | 0 | 0 | 4 | Moderate |
| **Garg et al.** | 1 | 0 | 1 | 0 | 0 | 0 | 0 | 0 | 0 | 2 | Low |
| **Gözen et al.** | 1 | 0 | 1 | 0 | 0 | 0 | 1 | 0 | 0 | 3 | Low |
| **Kumar et al.** | 0 | 0 | 1 | 0 | 0 | 0 | 1 | 0 | 0 | 2 | Low |
| **Petersen et al.** | 0 | 0 | 1 | 0 | 0 | 0 | 1 | 0 | 0 | 2 | Low |
| **Bulğurcu et al.** | 0 | 0 | 1 | 0 | 0 | 0 | 1 | 1 | 0 | 3 | Low |
| **Mazzatenta et al.** | 1 | 0 | 1 | 0 | 0 | 0 | 1 | 0 | 0 | 3 | Low |
| **Anna et al.** | 1 | 0 | 1 | 0 | 0 | 0 | 0 | 0 | 0 | 2 | Low |
| **Sayin et al. 2** | 1 | 0 | 1 | 0 | 1 | 0 | 1 | 0 | 0 | 4 | Moderate |
| **Staven at al.** | 1 | 0 | 1 | 0 | 0 | 0 | 1 | 0 | 0 | 3 | Low |
| **Ramteke et al.** | 1 | 0 | 1 | 1 | 0 | 0 | 1 | 0 | 0 | 4 | Moderate |
| **Lechner et al.** | 1 | 0 | 1 | 0 | 1 | 0 | 1 | 0 | 0 | 4 | Moderate |
| **Conchiero-Guisan et al.** | 1 | 0 | 1 | 0 | 0 | 0 | 1 | 1 | 0 | 4 | Moderate |
| **Horvath et al.** | 1 | 0 | 1 | 0 | 0 | 0 | 1 | 0 | 0 | 3 | Low |
| **Bastiani et al.** | 1 | 0 | 1 | 0 | 1 | 0 | 1 | 0 | 0 | 4 | Moderate |
| **Elimian et .** | 1 | 0 | 1 | 1 | 0 | 0 | 1 | 0 | 0 | 4 | Moderate |
| **Adamczyk et al.** | 1 | 1 | 1 | 1 | 0 | 0 | 1 | 0 | 0 | 5 | Moderate |
| **Lagi et al.** | 1 | 0 | 1 | 1 | 1 | 0 | 1 | 0 | 0 | 5 | Moderate |
| **Lapostolle et al.** | 1 | 0 | 1 | 0 | 0 | 0 | 1 | 0 | 0 | 3 | Low |
| **Zayet at al.** | 1 | 0 | 1 | 0 | 1 | 0 | 1 | 0 | 0 | 4 | Moderate |
| **Zou et al.** | 1 | 0 | 1 | 0 | 0 | 0 | 1 | 0 | 0 | 3 | Low |
| **Bianco et al.** | 1 | 0 | 1 | 0 | 0 | 0 | 1 | 0 | 0 | 3 | Low |
| **Andrews et al.** | 1 | 0 | 1 | 0 | 0 | 0 | 1 | 0 | 0 | 3 | Low |
| **Kavaz et al.** | 1 | 0 | 1 | 1 | 1 | 0 | 1 | 0 | 0 | 5 | Moderate |
| **Sahoo et al.** | 1 | 0 | 1 | 1 | 1 | 0 | 1 | 1 | 0 | 6 | Moderate |
| **Lechien & Cabaraux et al** | 1 | 0 | 1 | 0 | 0 | 0 | 0 | 0 | 0 | 2 | Low |
| **Lechien & Chiesa‐Estomba et al. 2** | 1 | 0 | 1 | 0 | 0 | 0 | 0 | 0 | 0 | 2 | Low |
| **Fisher et al.** | 1 | 0 | 1 | 0 | 0 | 0 | 1 | 0 | 0 | 3 | Low |
| **Niklassen et al.** | 1 | 0 | 1 | 0 | 0 | 0 | 0 | 1 | 0 | 3 | Low |
| **Nishanth et al.** | 1 | 0 | 1 | 0 | 1 | 0 | 1 | 0 | 0 | 4 | Moderate |
| **Song et al. 2** | 1 | 0 | 0 | 0 | 0 | 0 | 1 | 0 | 0 | 2 | Low |
| **Polat et al.** | 0 | 0 | 1 | 0 | 0 | 0 | 1 | 0 | 0 | 2 | Low |
| **Arslan et al.** | 1 | 0 | 1 | 0 | 1 | 0 | 1 | 0 | 0 | 4 | Moderate |
| **Yadav et al.** | 1 | 0 | 1 | 0 | 1 | 0 | 1 | 0 | 0 | 4 | Moderate |
| **Salcan et al.** | 1 | 0 | 1 | 0 | 0 | 0 | 1 | 0 | 0 | 3 | Low |
| **Amerigo et al.** | 1 | 0 | 1 | 0 | 1 | 0 | 1 | 0 | 0 | 4 | Moderate |
| **Fistera et al.** | 0 | 0 | 1 | 1 | 0 | 0 | 1 | 0 | 0 | 3 | Low |
| **Pinna et el.** | 1 | 0 | 1 | 0 | 0 | 0 | 0 | 0 | 0 | 2 | Low |
| **Liang et al.** | 1 | 0 | 1 | 0 | 0 | 0 | 1 | 0 | 0 | 3 | Low |
| **Magnavita et al.** | 1 | 0 | 1 | 0 | 1 | 0 | 1 | 0 | 0 | 4 | Moderate |
| **La Torre et al.** | 1 | 0 | 1 | 0 | 1 | 0 | 1 | 0 | 0 | 4 | Moderate |
| **Barillari et al.** | 1 | 0 | 1 | 0 | 1 | 0 | 1 | 0 | 0 | 4 | Moderate |
| **Singer-Cornelius et al.** | 1 | 0 | 1 | 0 | 0 | 0 | 1 | 0 | 0 | 3 | Low |
| **Kumar et al. 2** | 1 | 0 | 1 | 0 | 1 | 0 | 1 | 0 | 0 | 4 | Moderate |
| **Morlock et al.** | 0 | 0 | 1 | 0 | 0 | 0 | 1 | 0 | 0 | 2 | Low |
| **Thakur et al.** | 1 | 0 | 1 | 0 | 0 | 0 | 0 | 0 | 0 | 2 | Low |
| **Printza et al.** | 1 | 0 | 1 | 0 | 0 | 0 | 1 | 0 | 0 | 3 | Low |
| **Tsivgoulis et al.** | 1 | 0 | 1 | 0 | 0 | 0 | 1 | 0 | 0 | 3 | Low |
| **Karadas et al.** | 1 | 0 | 1 | 0 | 0 | 0 | 0 | 0 | 0 | 2 | Low |
| **Bagnasco et al.** | 1 | 0 | 1 | 0 | 0 | 0 | 1 | 0 | 0 | 3 | Low |
| **Ozcan et al.** | 1 | 0 | 1 | 0 | 0 | 0 | 1 | 0 | 0 | 3 | Low |
| **Akinbami et al.** | 1 | 0 | 1 | 0 | 0 | 0 | 1 | 0 | 0 | 3 | Low |
| **Ninchritz-Becerra et al.** | 1 | 0 | 1 | 0 | 0 | 0 | 1 | 0 | 0 | 3 | Low |
| **AlShakhs et al.** | 1 | 0 | 1 | 0 | 0 | 0 | 1 | 0 | 0 | 3 | Low |
| **Mannan et al.** | 1 | 0 | 1 | 0 | 0 | 0 | 1 | 0 | 0 | 3 | Low |
| **Cellejon-Leblic et al.** | 1 | 0 | 1 | 0 | 0 | 0 | 1 | 0 | 0 | 3 | Low |
| **Song et al.** | 1 | 0 | 1 | 0 | 1 | 0 | 1 | 0 | 0 | 4 | Moderate |
| **O'Sullivan et al.** | 1 | 0 | 1 | 0 | 1 | 0 | 1 | 0 | 0 | 4 | Moderate |
| **Savtale et al.** | 1 | 0 | 1 | 0 | 1 | 0 | 1 | 0 | 0 | 4 | Moderate |
| **Islek at el.** | 1 | 0 | 1 | 0 | 0 | 0 | 1 | 0 | 0 | 3 | Low |
| **Gupta et al.** | 1 | 0 | 1 | 0 | 0 | 0 | 1 | 0 | 0 | 3 | Low |
| **Cao et al.** | 1 | 0 | 1 | 0 | 0 | 0 | 0 | 0 | 0 | 2 | Low |
| **Jethani et al.** | 1 | 0 | 1 | 0 | 0 | 0 | 0 | 0 | 0 | 2 | Low |
| **Lombardi et al. 2** | 1 | 0 | 1 | 0 | 1 | 0 | 1 | 0 | 0 | 4 | Moderate |
| **Makaronidis** | 1 | 0 | 1 | 0 | 0 | 0 | 1 | 0 | 0 | 3 | Low |
| **Monti et al.** | 1 | 0 | 1 | 0 | 0 | 0 | 1 | 0 | 0 | 3 | Low |
| **Oda et al.** | 1 | 0 | 1 | 0 | 0 | 0 | 0 | 0 | 0 | 2 | Low |
| **Lampl et al.** | 1 | 0 | 1 | 0 | 1 | 0 | 1 | 0 | 0 | 4 | Moderate |
| **O'Keefe et al.** | 0 | 0 | 1 | 0 | 0 | 0 | 1 | 1 | 0 | 3 | Low |
| **Asai et al.** | 1 | 0 | 1 | 0 | 1 | 0 | 1 | 1 | 0 | 5 | Moderate |
| **Martinez et al.** | 1 | 0 | 1 | 0 | 0 | 0 | 1 | 0 | 0 | 3 | Low |
| **Lee et al. 3** | 1 | 0 | 1 | 0 | 0 | 0 | 1 | 0 | 0 | 3 | Low |
| **Fleischer et al.** | 1 | 0 | 1 | 0 | 0 | 0 | 1 | 1 | 0 | 4 | Moderate |
| **Sun et al.** | 1 | 0 | 1 | 0 | 0 | 0 | 1 | 1 | 0 | 4 | Moderate |
| **Gibbons et al.** | 1 | 0 | 1 | 1 | 0 | 0 | 1 | 0 | 0 | 4 | Moderate |
| **Sbrana et al.** | 1 | 0 | 0 | 0 | 0 | 0 | 1 | 0 | 0 | 2 | Low |
| **Omezli & Torul** | 1 | 0 | 1 | 0 | 0 | 0 | 1 | 0 | 0 | 3 | Low |
| **Ladoire et al.** | 0 | 0 | 1 | 1 | 0 | 0 | 1 | 0 | 0 | 3 | Low |
| **Boscolo-Rizzo et al.** | 1 | 0 | 1 | 0 | 0 | 0 | 1 | 0 | 0 | 3 | Low |
| **Schwab et al.** | 1 | 0 | 1 | 1 | 0 | 0 | 0 | 0 | 0 | 3 | Low |
| **Rodebaugh et al.** | 1 | 0 | 1 | 0 | 0 | 0 | 1 | 0 | 0 | 3 | Low |
| **Zejda et al.** | 1 | 0 | 0 | 0 | 0 | 0 | 1 | 1 | 0 | 3 | Low |
| **Printza et al. 2** | 1 | 0 | 0 | 1 | 0 | 0 | 1 | 0 | 0 | 3 | Low |
| **Trachootham et al.** | 1 | 0 | 1 | 0 | 0 | 0 | 1 | 0 | 0 | 3 | Low |
| **Sehanobish et al.** | 0 | 0 | 0 | 0 | 0 | 0 | 1 | 0 | 0 | 1 | Low |
| **Gurrola et al.** | 1 | 0 | 0 | 1 | 0 | 0 | 1 | 0 | 0 | 3 | Low |
| **Hijazi et al.** | 1 | 0 | 1 | 0 | 0 | 0 | 1 | 0 | 0 | 3 | Low |
| **Besli et al.** | 1 | 0 | 1 | 0 | 1 | 0 | 1 | 0 | 0 | 4 | Moderate |
| **Dini et al.** | 1 | 0 | 1 | 0 | 1 | 0 | 1 | 0 | 0 | 4 | Moderate |
| **Gonzalez et al.** | 1 | 0 | 1 | 0 | 1 | 0 | 1 | 1 | 0 | 5 | Moderate |
| **Chung et al.** | 0 | 0 | 1 | 0 | 0 | 0 | 1 | 0 | 0 | 2 | Low |
| **Kandakure et al.** | 1 | 0 | 1 | 0 | 1 | 0 | 0 | 0 | 0 | 3 | Low |
| **Dixon et al.** | 1 | 0 | 1 | 0 | 0 | 0 | 1 | 0 | 0 | 3 | Low |
| **Boudjema et al.** | 1 | 0 | 1 | 0 | 0 | 0 | 1 | 0 | 0 | 3 | Low |
| **Galluzzi et al.** | 1 | 0 | 1 | 0 | 0 | 0 | 1 | 0 | 0 | 3 | Low |
| **Zifko et al.** | 1 | 0 | 1 | 0 | 0 | 0 | 1 | 0 | 0 | 3 | Low |
| **Amano et al.** | 1 | 0 | 1 | 0 | 1 | 0 | 1 | 1 | 0 | 5 | Moderate |
| **Ismail et al.** | 1 | 0 | 1 | 0 | 1 | 0 | 1 | 0 | 0 | 4 | Moderate |
| **Garcia et al.** | 1 | 0 | 1 | 0 | 1 | 0 | 1 | 0 | 0 | 4 | Moderate |
| **Tarifi et al.** | 1 | 0 | 1 | 0 | 0 | 0 | 1 | 0 | 0 | 3 | Low |
| **van der Besselaar** | 0 | 0 | 1 | 0 | 0 | 0 | 0 | 0 | 0 | 1 | Low |
| **Gianola et al.** | 1 | 0 | 1 | 0 | 1 | 0 | 1 | 0 | 0 | 4 | Moderate |
| **Teklu et al.** | 1 | 0 | 1 | 1 | 0 | 0 | 1 | 0 | 0 | 4 | Moderate |
| **El Kady et al.** | 1 | 0 | 1 | 0 | 1 | 0 | 1 | 0 | 0 | 4 | Moderate |
| **Gupta et al. 2** | 1 | 0 | 0 | 0 | 0 | 0 | 1 | 0 | 0 | 2 | Low |
| **Alharbi et al.** | 0 | 0 | 1 | 0 | 0 | 0 | 1 | 0 | 0 | 2 | Low |
| **Karaarslan et al.** | 1 | 0 | 1 | 0 | 1 | 0 | 1 | 0 | 0 | 4 | Moderate |
| **Breyer et al.** | 1 | 0 | 1 | 0 | 0 | 0 | 1 | 0 | 0 | 3 | Low |
| **Parcha et al.** | 1 | 0 | 1 | 0 | 0 | 0 | 1 | 0 | 0 | 3 | Low |
| **Rousseau et al.** | 1 | 0 | 1 | 0 | 0 | 0 | 1 | 0 | 0 | 3 | Low |
| **Le Bon et al. 3** | 0 | 0 | 1 | 0 | 1 | 0 | 1 | 0 | 0 | 3 | Low |
| **Kamel et al.** | 1 | 0 | 1 | 0 | 1 | 0 | 1 | 0 | 0 | 4 | Moderate |
| **Ashrafi et al.** | 1 | 0 | 1 | 0 | 1 | 0 | 1 | 0 | 0 | 4 | Moderate |
| **Wierdsma et al.** | 1 | 0 | 1 | 0 | 0 | 0 | 1 | 1 | 0 | 4 | Moderate |
| **Moeller et al.** | 1 | 0 | 0 | 0 | 0 | 0 | 1 | 0 | 0 | 2 | Low |
| **de Torres et al.** | 1 | 0 | 1 | 0 | 0 | 0 | 1 | 0 | 0 | 3 | Low |
| **Noviello et al.** | 1 | 0 | 1 | 1 | 0 | 0 | 1 | 0 | 0 | 4 | Moderate |

Source: From Hoy et al. (2012), as adapted by Tong et al. (2020).

^A^ Questions were scored as 0 = no, 1 = yes and comprised the following:

Q1: Was the study’s target population a close representation of the national population in relation to relevant variables, e.g., age, sex, occupation?

Q2: Was the sampling frame a true or close representation of the target population?

Q3: Was some form of random selection used to select the sample, or was a census undertaken?

Q4: Was the likelihood of nonresponse bias minimal?

Q5: Were data collected directly from the subjects (as opposed to a proxy)?

Q6: Was an acceptable case definition used in the study?

Q7: Was the study instrument that measured the parameter of interest (e.g., prevalence of low back pain) shown to have reliability and validity (if necessary)?

Q8: Was the same mode of data collection used for all subjects?

Q9: Were the numerator(s) and denominator(s) for the parameter of interest appropriate?

^B^ Risk-of-bias categories were as follows: 0–3, low; 4-6, moderate; 7-9, high.

**S3: Supplementary Table. Random-effect estimate of taste classification on COVID-19 taste loss prevalence using generalized linear mixed models.**

| **Measure** | **k** | **Proportion** | **95% CI** | **Q** | **I^2^** | **tau^2^** |
| --- | --- | --- | --- | --- | --- | --- |
| Taste Only; Taste and Smell | 18 | 0.3961 | 0.2642-0.5451 | 1370.8 | 98.80% | 1.4082 |
| Smell AND/OR Taste | 19 | 0.3724 | 0.2301-0.5409 | 716.7 | 97.50% | 1.9376 |
| Taste Only | 153 | 0.3902 | 0.3454-0.4370 | 9033.47 | 98.30% | 1.3989 |
| Smell AND Taste | 15 | 0.238 | 0.1320-0.3907 | 11454.02 | 99.90% | 1.6249 |
| Smell OR Taste | 36 | 0.3266 | 0.2393-0.4278 | 1993.87 | 98.20% | 1.5649 |
